# Validation of a Visual Midline Gauge: A Cross-Sectional Study

**DOI:** 10.1101/2023.04.25.23289127

**Authors:** Amritha Stalin, Ran Ding, Susan J. Leat, Ohwod Binhilabi, Tammy Labreche

## Abstract

**Background:** This study standardized the parameters of a novel visual midline gauge and presents normative data, which can be used by optometrists in the assessment of visual midline.

**Methods:** Ninety-three participants from three age groups (18 to 44, 45 to 64, and > 65 years) without history of significant neurological or ocular problems were recruited in Waterloo, Canada and Hong Kong. In experiment 1, the perceived horizontal and vertical visual midline was measured using the gauge for 2 speeds and 2 repositioning methods. In experiment 2, the perceived midline was measured for three different distances (25, 50 and 100 cm) using a target speed and repositioning method chosen from the first experiment results. Since there was no significant difference between the two sites in any of the measures, data were combined for analysis.

**Results:** For experiment 1, linear mixed models showed no effect of age, speed or repositioning method and no interaction (p>0.05) for the perceived midline position (p>0.05). In experiment 2, there was no significant effect of age or distance and no interaction effects on the perceived horizontal and vertical visual midline position using the chosen speed and method (2.3 degrees per second, adjustment method). Normative data (mean and 95% ranges of the perceived visual midline for control participants) was tabulated. The measurements were found to be repeatable. A few participants were found to have significant midline shifts.

**Conclusion:** This study shows that the measurement of midline is tolerant of differences of target speed, testing method, test distance, and age group of the participants and that the measurements using the visual midline gauge are repeatable. It is possible for even individuals without a history of neurological or ocular disorders to have significant shifts in their perceived visual midline.

## INTRODUCTION

Cerebrovascular disease is the sixth most common global cause of chronic disability.^1,2^ According to the Canadian Heart and Stroke Foundation, the number of cerebrovascular accidents (CVAs or stroke) in Canada has increased^3^ and this is also the case in the UK^4^, US^5,6^, developing countries^7^ and worldwide.^1,2^ The incidence of stroke increased by 74% between 1981 to 2004 in the UK for people older than 75 years old.^4^ Between 550,000 to 795,000 people experience a stroke each year in the United States ^5,6^, and around 400,000 Canadians are living with post-stroke disabilities.^3^ There is also an increase in the number of people surviving a stroke and therefore living with the consequences.^8^ In the United States, 70% of stroke survivors receive rehabilitation services through either hospital-based inpatient, nursing, or home rehabilitation care outpatient facilities.^5^ Approximately 10% of individuals are admitted to a long term care facility after a stroke and only a few recover completely.^3,9–11^ It is estimated that all stroke survivors have some degree of visual disorder, 60% having minor visual impairment and 40% surviving with moderate to severe impairments.^3,9,10^

Within the optometric profession, there is an increased interest in rehabilitation to enhance the independence, function, safety, and health of an individual and to optimise aging following a neurological event. After a CVA, individuals may have a wide range of visual sequelae, including impaired visual acuity, binocular vision disorders, visual field deficits, and visual perceptual difficulties.^12,13^ Up to 70% of stroke patients report visual field defects.^10^ Reduced visual acuity post-stroke (less than 0.50 logMAR [6/18] corrected) was documented in 13% of individuals for both near and distance vision.^12^ Ocular motility deficits are present in 68.4% of individuals with stroke.^12^ Perceptual changes include visual neglect (unilateral spatial inattention), and visual midline shift, in addition to others.

Visual neglect is a neurological disorder which occurs after a brain lesion and can result in loss of visual attention of one side of visual space.^14–17^ Neglect can impact different personal spaces (personal space, intrapersonal-space, and external space) and is the most common perceptual change post-stroke.^12^

Abnormal egocentric localization or visual midline shift (VMS) is a condition in which an individual experiences a shift in their visual midline, defined as the point which the individual perceives as being straight ahead or “in front of their nose”.^9,10,18^ The midline can be shifted laterally or vertically so that a target will be perceived as straight ahead when, for example, it is presented in front of the left eye rather than the true inter-ocular centre.^18,19^ The prevalence of the visual midline shift following CVA has not been determined. Visual midline shift may result in postural changes and subsequent issues with balance and safety, the resultant condition being called visual midline shift syndrome.^3,9,10,13,18^

The pathophysiology of visual midline shift is uncertain and there is discrepancy in the terminology used. It has been proposed that a CVA may affect a specific area of the feed-forward dorsal visual pathway within the right inferior or posterior parietal lobe of the brain resulting in visual neglect.^20^ Neglect of one hemisphere may result in a change of visual midline, often referred to as an altered subjective-straight ahead.^18,20^ Padula et al.^18^ proposed that as a result of hemianopia following a CVA, VMS develops as a result of lack of information to the retinotectal pathway. This visual pathway travels from the retina through the superior colliculus to the pulvinar nucleus in the midbrain instead of through the lateral geniculate nucleus and onto the primary visual cortex.^18^ Information that travels though this pathway combines with information from different systems such as the vestibular, proprioceptive, and kinesthetic which are responsible for the awareness of self-space by an individual.^18^ Therefore, lack of information provided to that pathway causes the brain to attempt to re-create a balance which results in a postural lean away from the affected visual space disturbing posture and balance.^18^

There has been little research regarding VMS, leaving many questions to be answered in terms of the epidemiology, pathophysiology, and evidence-based treatment modalities. This contrasts with the on-going and considerable research into visual neglect which may accompany VMS. Prisms have been suggested as a rehabilitation intervention for patients with VMS to correct posture and balance, improve quality of life following a stroke and possibly decrease the incidence of falls.^13,21,22^ However, there are no clear guidelines regarding duration of wear or amount of prism. ^22,23^ There is also insufficient evidence regarding accurate assessment of VMS, which is necessary for any study protocol or clinical management.

At present there is no validated clinical method of measuring the visual midline. Clinically, the visual midline is currently measured manually. The patient is asked to visually track a “wand” (a hand-held target) moved laterally or vertically in front of the individual to find the subjective visual midline point. Padula et al. ^18^ used a 30 cm long wand, the tip of which is the target, presented against a blank wall at approximately 45 cm from the patient. The recommended speed of the target was 4 cm per second. Various other methods have been described. For example, Houston^20^ called the visual midline the visual eye-head egocentric midline and created an apparatus to standardize measurements. Kapoor et al.^13^ used a laser pointer, viewed through a mirror while Kuhn et al.^23^ described a method to measure the midline (called the subjective straight ahead) with a small red spot presented in a perimeter.^23^

The purpose of this study is to standardize the parameters of a visual midline gauge using participants with unimpaired vision and no history of stroke or other neurological disorders.^18^ The information gained in this study will be used to develop an accurate assessment tool for use with post-stroke patients and to gather normative data against which the visual midline of post-CVA patients can be compared.

## METHODS

The Visual Midline (VM) gauge consists of a metal bar against a solid black background. The bar and the background are both the same colour black. A white sphere of 13 mm size is situated just behind the bar and can be moved at different speeds (1.15 or 2.3 degrees per second). The target moves mechanically and can be manoeuvred using a control box and push button. The bar has a millimetre scale for the examiner to measure the position of the target, which is not visible to the participant (Figure 1).

**Figure 1:**
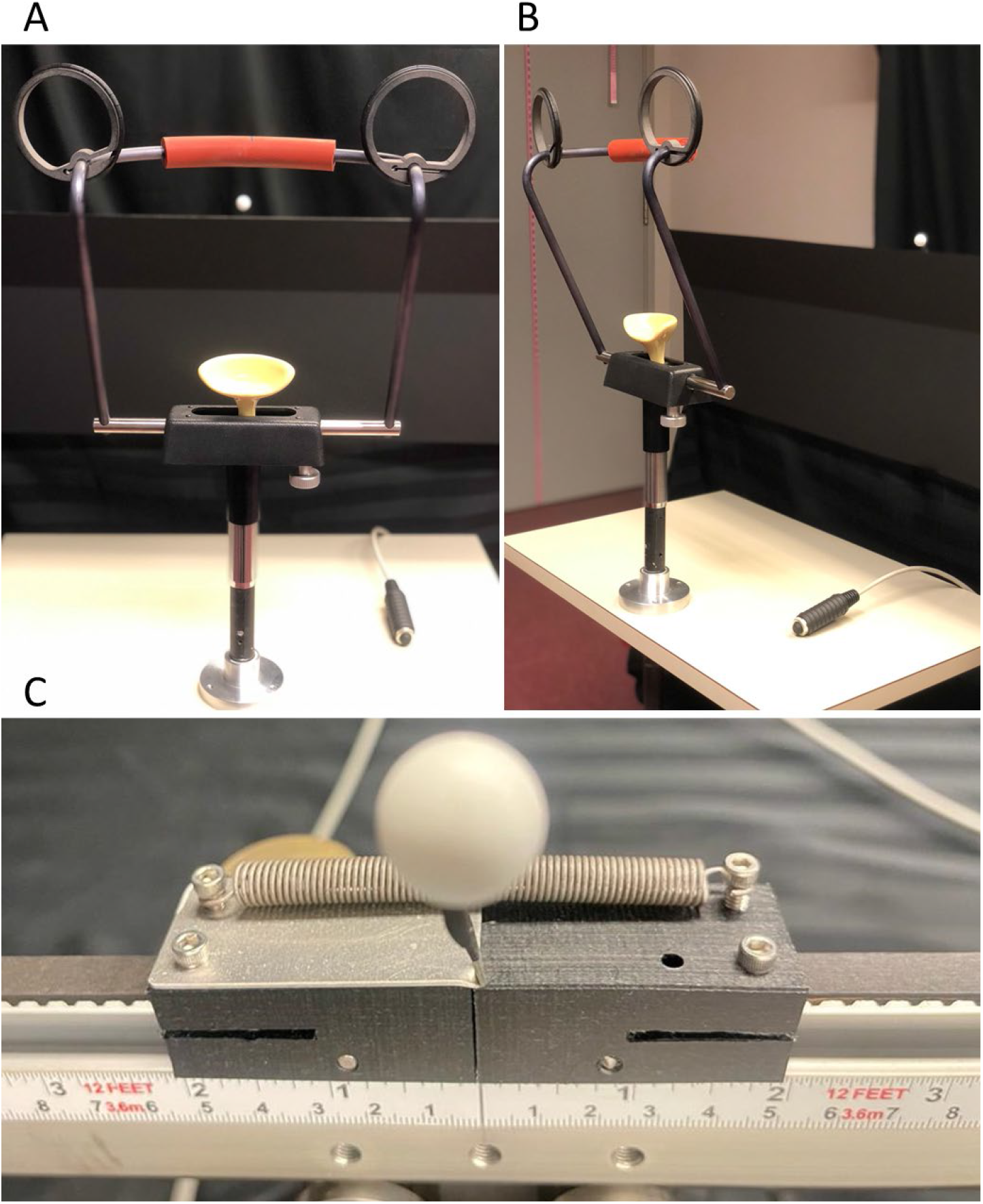
Post stoke visual midline gauge, A & B: Table with chin rest, and buzzer. C: Experimenter’s view from the back showing the numbered scale and target.

This cross-sectional study received clearance through a University of Waterloo Research Ethics Board and a Hong Kong Polytechnic University (PolyU) Institutional Review Board and adhered to the principles of the Declaration of Helsinki. Participants were recruited from among the graduate students, staff, faculty, and their families as well as the School of Optometry Clinics at the University of Waterloo (UW) School of Optometry and Vision Sciences (UW) and the PolyU from January 2022 to September 2022. Snowball recruiting was also used.

A preliminary phone screening interview was conducted to ensure that potential participants met the ocular and health history inclusion and exclusion criteria. Inclusion criteria were; either gender, corrected visual acuity of 0.1 logMAR (6/7.5) or better in each eye. Participants were excluded if they met any of the following criteria: more than one-line intra-ocular difference in visual acuity (VA), strabismus, significant previous ocular or health history, neurological disorders such as Parkinson’s, multiple sclerosis, cerebellar dysfunction, vestibular dysfunction, cerebral palsy, dizziness/vertigo, a diagnosis of dementia or cognitive impairment, previous history of stroke or any medication that would affect balance. In addition, the phone interview included questions on handedness (right, left or ambidextrous) and history of falls.

Fifty-four and 39 participants were recruited from UW and PolyU respectively, in three age groups; 18 to 44, 45 to 64, and above 65 years (See Table 1).

**Table 1:** Demographic data of all participants.

| Age group | UW | PolyU |
| --- | --- | --- |
| 18 to 44 years | 13 women & 7 men<br>29.4±6.7 years (18 to 44) | 9 women & 5 men<br>25.3±6.5 years (19 to 42) |
| 45 to 64 years | 8 women & 9 men<br>54.9±6.8 years (45 to 64) | 5 women & 7 men<br>59.7±5.3 years (47 to 64) |
| 65 years and above | 9 women & 8 men<br>72.0±5.3 years (65 to 84) | 9 women & 4 men<br>69.5±3.5 years (65 to 77) |

Participants attended for two visits. During the first visit, participant’s VA was measured at 4m using a different Early Treatment Diabetic Retinopathy Studies chart for each eye. A per-letter scoring system was used for calculating the VA, which was recorded in logMAR units.^24^ Ocular alignment was assessed with the unilateral cover test for distance and near using an accommodative target, and visual field loss (VF) was assessed using a hand-held arc perimeter with a 13 mm diameter white target and recorded manually on Goldmann VF recording sheets. After confirming the eligibility using these tests, the VM assessments were conducted.

Visual midlines were assessed using the clinical method and the VM gauge. During the clinical method,^18^ participants were asked to visually track a hand-held 13 mm diameter sphere target on a 30 cm long wand that was moved laterally or vertically in front of their face at a distance of 50 cm. The speed of the target was approximately 3-4 cm per second. Participants were asked to indicate when the target was perceived to be in their midline. For horizontal measurements they were asked to indicate when the target was in line with the tip of their nose and for vertical measurements, they were asked to respond when it was level with their eyes. The examiner subjectively estimated the VM from their intra-ocular midline in centimeters based on the participants response. Two trials were undertaken from each direction.

At the first visit (Experiment 1), horizontal and vertical VM was measured using the VM gauge at a 50 cm distance, with 2 speeds (1.15 or 2.3 degrees per second) and two methods of repositioning the target for accuracy (adjustment and repeated). Horizontal and vertical VM was also measured with the clinical method (see description above). At the second visit (Experiment 2), vertical and horizontal VM was measured for 25, 50 and 100 cm using the speed and method of repositioning chosen based on the data analysis of all the first visit data. VM with the clinical method was repeated.

During the measurement with the VM gauge, the participant was seated at the specified distance in front of the midline gauge. A headrest table was used to stabilize head position, adjusted for the target distance such that their inter-ocular midpoint was in line with the ‘0’ midpoint on the device horizontally and level with the centre of the spherical target vertically. The room lights were fully on.

Before each measurement, the target was placed in a position remote from the anticipated midline of the participant on alternating sides (left or right) and moved towards the expected midline position. The ‘0’ position on the VM gauge was not visible to the participant. The participant was instructed to visually track the target with their eyes. The participant responded with a push button when they perceived the target to be aligned with their visual midline, which stopped the target movement. For horizontal measurements they were asked to press the push button when the target was in line with the tip of their nose. For vertical measurements, they were asked to respond when it was level with their eyes. Five trials were undertaken from each direction. Participants were allowed to reposition the target position after each trial if they did not believe it was in line with their midline. Two different methods were used in visit 1 for this correction. In the adjustment method, participants were allowed to ask the examiner to restart the trial and the participant was able to adjust the position of the target from where it stopped initially using the push button. In the repeated method, the target was moved back to the initial position (similar to when the trial started), and the trial was repeated until the participant confirmed that the final target position was in alignment with their perceived midline. This repeated method required 3 to 4 remeasurements for some of the participants.

### Experiment 1

The parameters for the VM gauge measurement in visit 1 (vertical/horizontal, two speeds, and two methods of adjustment) were varied in a counter-balanced order between participants to control for fatigue and practice effects. Two trials were performed in each direction to demonstrate the task, and the results of these trials were not recorded. Following this, the measurements were repeated five times from each direction resulting in a total of 80 trials for each participant during the first visit. Breaks were provided in between the conditions to reduce fatigue and to maintain attention.

### Experiment 2

During visit 2, VM testing was conducted with the target speed (2.3 degrees per second) and the adjustment method of repositioning determined from the results of the first experiment. In experiment 2, the horizontal and vertical midlines were measured at 50 and 100 cm distances, corresponding to intrapersonal-space and external space), but only horizontal measurements were possible at 25 cm (personal space) due to mechanical constraints. Once again, the parameters were varied in a counter-balanced order and five measurements were taken from left, right, up and down starting points.

#### Analysis

Jamovi (version 2.3.18.0) software and Microsoft office Excel (version 16.55) were used to analyze the data. Descriptive statistics were examined to ensure that data met statistical test assumptions and Shapiro-Wilk test was used to assess normality. A Mann-Whitney U-test was conducted to compare the VM data from UW and PolyU since the examiners were different at each location. There was no significant difference (p>0.05) in any of the measures, therefore, data from both centres were combined for all further analysis.

Since the data were not normally distributed, Wilcoxon signed-rank tests were used to assess the effect of test direction (right vs. left and up vs. down) at all distances for both visits. Linear mixed models for repeated measures were used to analyze the impact of target speed, method of repositioning, and age group of participants on the VM at first visit for the horizontal and vertical directions. Linear mixed models were also used to analyze the effect of testing distance and age group of participants on the VM at second visit for the horizontal and vertical visual midline.

A Wilcoxon W test was used for each participant to check the study assumption that these normal observers did not have a significant midline shift, i.e., whether their perceived midline was significantly different from zero. Since many t-tests were undertaken (for each distance, vertical and horizontal) a Bonferroni correction was used and the usual p value of <0.05 was changed to <0.05/n = <0.00056 for significance. One sample t-tests were also performed for each distance for both horizontal and vertical measures for each age group to determine if the average position of the midline was centred on zero.

Clinically, it is usual to perform two measurements from each side.^18^ So normal data for use in clinical settings were calculated as the mean of the first two measurements from each direction and 1.96 x SD (95% range). All other analysis was based on the full data set of 5 readings from each direction) parameters.

Repeatability between visit 1 and 2 was assessed using Bland Altman plots, bias, limits of agreement for repeatability, and correlation coefficients.

## RESULTS

### Experiment 1

#### Effect of direction

The results of Wilcoxon signed-rank tests showed significant differences between the visual midlines measured with different starting directions (i.e., when the target moved from right vs. left, and up vs. down). Specifically, the visual midlines were found to be shifted towards the direction from where the target started (Figure 2). Hence, the 10 measurements for each person (5 from each direction) were paired and analyzed as one measurement for all further analysis. For example, the first measurement from the right was paired with the first measurement from the left as one data point. This is similar to how the tests would be conducted clinically^18^.

**Figure 2.**
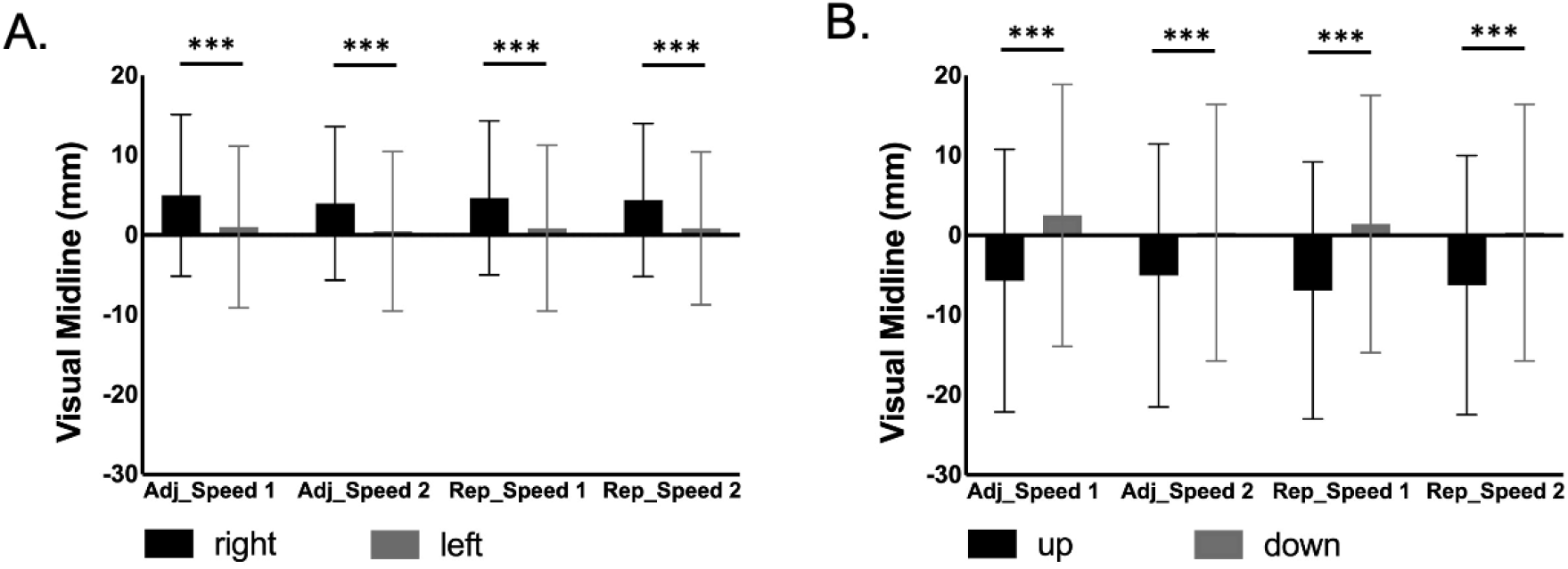
The effect of direction. A. The mean and standard deviation of the visual midline for the first visit, horizontal direction (right vs. left) for 2 different speeds and 2 repositioning methods. For each participant, the average of the 10 measures from the right and left were calculated. Positive results indicate rightward shifts. B. The mean and standard deviation of the visual midline shift for the first visit, vertical direction (up vs. down) for 2 different speeds and 2 repositioning methods. For each participant, the average of the 10 measures from up and down are plotted. Positive results indicate downward shifts. “adj” = adjustment method, “rep” = repeated method. * *p* < 0.05, ** *p* < 0.01, *** *p* < 0.001

#### Effect of method and target speed

The linear mixed model included fixed effects for age group, target speed, repositioning method as well as the interactions and a random intercept per participant. There was no significant effect of repositioning method, target speed, or age group on the position of visual midline for either horizontal or vertical directions (p>0.05). There were no significant interaction effects. To conduct the VM test more efficiently, the faster repositioning method (i.e., adjustment) and speed (i.e.,2.3 degrees/second) were chosen for the second visit.

### Experiment 2

The linear mixed model for the second visit included fixed effect for age group and distance as well as the interactions and a random intercept per subject. There was no significant effect of distance or age group for either horizontal or vertical direction (p < 0.05). There were no significant interaction effects.

#### Visual midline shifts in individuals

Wilcoxon W tests were conducted to assess for a significant VMS in each participant. Each participant’s average midline position was compared with zero, for the method of adjustment, speed of 2.3 degrees/s, and distance of 50cm. This showed that some individuals had significant horizontal and vertical visual midline shifts which remained significant after Bonferroni correction (modified p value was <0.05/91 = <0.00055). In age group 1, there were 2 and 3 participants who had horizontal and vertical VMSs, respectively. In age group 2, there were 4 and 7 participants who had horizontal and vertical VMSs, respectively. and in age group 3, there were 12 and 9 participants who had horizontal and vertical VMSs, respectively.

#### Repeatability

The first two measurements from each starting direction were used to assess the repeatability. The results of the Bland Altman plots, and the Spearman correlation analysis are shown in Figures 3 & 4. The Bland Altman plots, limits of agreement for repeatability, and correlation coefficients all showed good repeatability.

**Figure 3.**
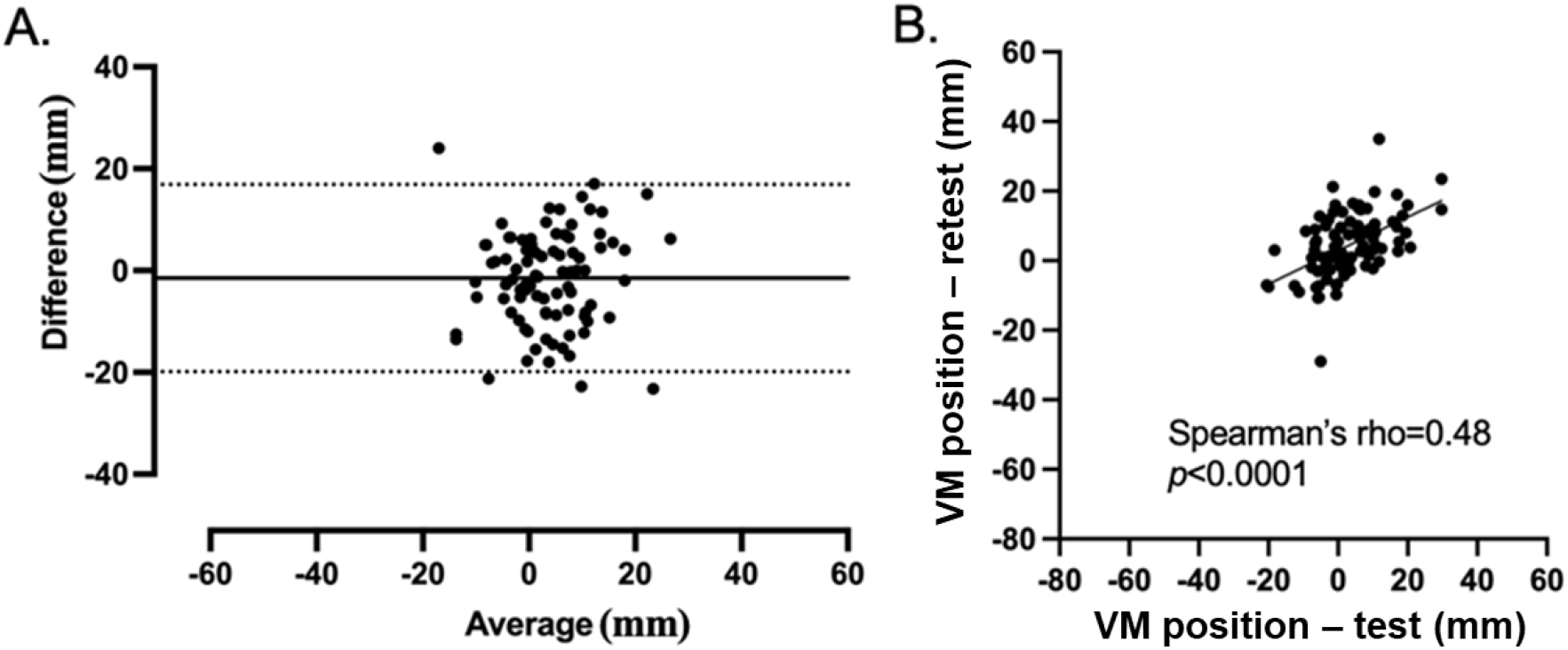
**A:** Bland Altman plots for test-retest repeatability of the difference between the first and second visit for the adjustment method, horizontal direction, 50 cm. The solid line indicates the average of the differences between the two visits. The dotted lines represent the upper and lower limits of agreement. **B:** Scattergrams of test and retest for the adjustment method, horizontal direction, 50cm.

**Figure 4.**
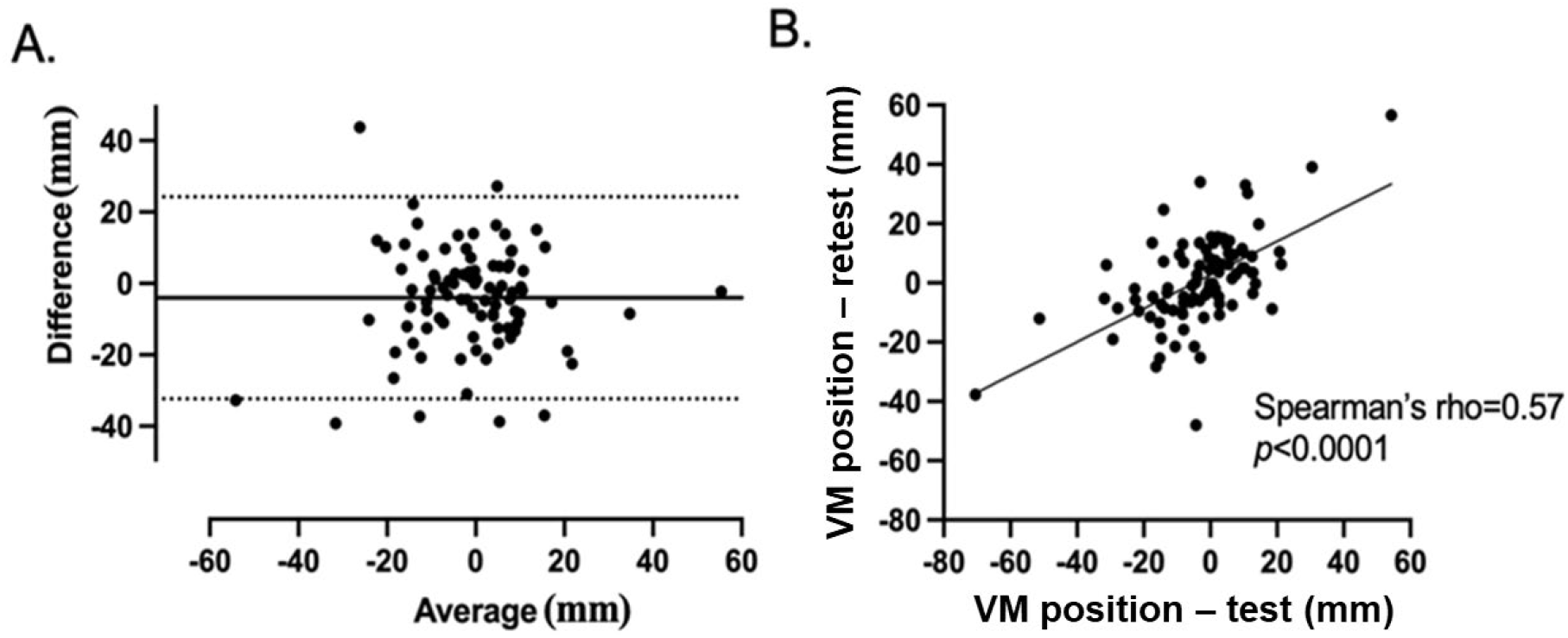
**A:** The result of the Bland Altman plots for test-retest repeatability of the difference between the first and second visit for the adjustment method, vertical direction, 50cm. The solid line indicates the average of the differences between the two visits. The dotted lines represent the upper and lower limits of agreement. **B:** Scattergrams of test and retest for the adjustment method, vertical direction, 50cm.

The results were similar when test-retest repeatability analysis was conducted with all 10 measurements (5 paired measurements).

#### Normative data

Because there was no effect of age or testing distance of 50 and 100 cm, normative data were calculated for all ages and 50 and 100 cm together. The data for 25cm were kept separate since measurements at 25 cm were only possible for the horizontal direction. Means and the 95% range of normal (1.96 x SD) are shown in Table 2.

**Table 2:** Normative data for all age groups for the horizontal and vertical midline positions in millimeters. Positive results indicate right/downward shifts.

|  | Mean<br>(mm) | Standard<br>deviation<br>(mm) | Minimum<br>(mm) | Maximum<br>(mm) | Range<br>(1.96 X SD) |  |
| --- | --- | --- | --- | --- | --- | --- |
| Horizontal<br>(25 cm) | 2.53 | 6.2 | -15.7 | 21.4 | -9.622 | 14.682 |
| Horizontal<br>(50&100 cm) | 1.4 | 7.94 | -31.3 | 23.9 | -14.1624 | 16.9624 |
| Vertical<br>(50&100 cm) | 1.15 | 10.5 | -21.6 | 50.6 | -19.43 | 21.73 |

## DISCUSSION

The purpose of the study was to develop and test a new device to measure the perceived visual midline position. This study investigated the effect of parameters such as target speed, repositioning method, testing distance, and age of participants in individuals without a history of significant neurological or ocular disorders. The study is important, as, although other authors have described various methods for measuring the midline, there are currently no published studies on the effect of the various parameters on the VM or normative data.

There are a wide variety of suggested ways to measure the perceived midline. Padula et al.^18^ described using one target size (30 cm length) and one speed (4 cm per second) at one distance (45 cm), but no explanation was provided on the basis of choosing these parameters. The portable method mentioned by Kapoor et al.^13^ used a laser pointer target moving slowly in front of the participant while the participant looked through goggles and saw a reflected mirror image in front of their eyes. A hand-held clicker was used to indicate when the image of the laser pointer was aligned with their midline. Some of the participants were instructed to look straight and not to follow the target and other participants received a demonstration of the true subjective straight-ahead position prior to the test.^13^ The method by Kuhn et al.^23^ for measuring the visual midline shift involved a red spot presented in the perimeter in different quadrants right, left, up and down and the participant was asked to verbally adjust the position of the red spot until it aligned with their subjective straight-ahead position. Houston^20^ measured the visual eye-head egocentric midline by asking the participant to respond when a 1cm, manually moved white target (3 degrees/second) was perceived at the center.^20^

In the first session, we found no significant effect of target speed and repositioning method. So, the fastest speed and testing method (adjustment) were chosen based on clinical considerations as it would make the testing more efficient in a clinical setting. This speed was slower than the one used by Padula et al.^18^, which was 5 degrees per second, but much faster than that by Kapoor et al.^13^, which was 0.5 degrees per second.

The effect of starting direction was analysed from the first visit results, and there was a significant effect of starting direction. Therefore, it is important in any clinical or experimental setting that the final determination is based on the average of an equal number of measurements from each direction, as is indeed clinical practice^18^.

In Experiment 2, the midline position was measured at three different distances to see whether the perceived midline changes in personal space, intrapersonal-space, and external space. This is important as one underlying pathophysiological theory is that an altered midline occurs as a result of neglect^25^, which may occur differently in these spaces. Only the horizontal direction assessment was possible at the personal space level due to physical constraints. Experiment 2 showed no significant differences in the position of the perceived midline, based on age and distance. Therefore, the normal data is presented together for all age groups, separately for 25 cm, and combined for 50 and 100 cm. Overall, VM measurements were not significantly different from zero.

In this study, we expected that people with normal vision and no history of neurological impairments would have no significant midline shift. However, there were a few participants who had significant VMS when the individual measurements were compared with zero. To the authors’ knowledge, this finding has not been reported before, but is important as not all midline shifts may be due to stroke and may be present prior to stroke.

Prisms are one suggested method to control and treat midline shifts by two approaches. Some have recommended the prescription of prisms in glasses for full-time wear^18^ and others have recommended prism adaptation while undergoing therapy as a management.^19^ In either case it would be expected that there should be some relationship between the degree of midline shift and the amount of prism used and so an accurate measure of the presence and degree of midline shift is desired, as can be provided by this gauge.

This is the first research study that has attempted to standardize the assessment of visual midline. The study has shown that the measurement of the perceived midline is relatively insensitive to the target speed and testing distances within the range that was tested. This is also the first study to provide normative data, against which post-stroke patients can be compared.

Futures studies are indicated to use the VM gauge to determine the range and potential causes of midline shifts in individuals without stroke, but who might have other ocular disorders. It is also important to determine the prevalence and natural history of midline shift in the post-stroke population. Secondary analysis of this data will be conducted to determine the associations of VMS with other vision and non-vision variables.

## Data Availability

Supporting data is not available in a repository as participants of this study did not give written consent for their data to be shared publicly.

## NON-STANDARD ABBREVIATIONS AND ACRONYMS

CVA: Cerebrovascular accidents
PolyU: Hong Kong Polytechnic University
UW: University of Waterloo
VA: visual acuity
VF: visual field loss
VMS: visual midline shift
VM: visual midline

## ACKNOWLEDGEMENTS

We would like to thank Dr. Henry Chan for supporting the project at Hong Kong Polytechnic University.

## REFERENCES

1. Ezejimofor MC, Chen YF, Kandala NB, et al. Stroke survivors in low-and middle-income countries: a meta-analysis of prevalence and secular trends. J Neurol Sci. 2016;364:68–76.

2. Akbarkhodjaeva ZA, Tursunov J. Prevalence of stroke in the world. Journal of Stroke and Cerebrovascular Diseases. 2017;26(4):913–914.

3. Heart and Stroke Foundation of Canada. Different Strokes Recovery triumphs and challenges at any age. 2017 Stroke Report.. Accessed November 8, 2022. https://www.heartandstroke.ca/-/media/pdf-files/canada/stroke-report/strokereport2017en.ashx?la=en&hash=67F86E4C3338D5A7FE7862EA5D0DD57CA8539847

4. Rothwell PM, Coull AJ, Giles MF, et al. Change in stroke incidence, mortality, case-fatality, severity, and risk factors in Oxfordshire, UK from 1981 to 2004 (Oxford Vascular Study). The Lancet. 2004;363(9425):1925–1933.

5. Gresham GE, Stason WB, Duncan PW. Post-Stroke Rehabilitation. Vol 95. Diane Publishing; 2004.

6. Fang MC, Perraillon MC, Ghosh K, Cutler DM, Rosen AB. Trends in stroke rates, risk, and outcomes in the United States, 1988 to 2008. Am J Med. 2014;127(7):608–615.

7. Benamer HTS, Grosset D. Stroke in Arab countries: a systematic literature review. J Neurol Sci. 2009;284(1-2):18–23.

8. Johnson CO, Nguyen M, Roth GA, et al. Global, regional, and national burden of stroke, 1990–2016: a systematic analysis for the Global Burden of Disease Study 2016. Lancet Neurol. 2019;18(5):439–458.

9. Goldstein LB, Adams R, Becker K, et al. Primary prevention of ischemic stroke: a statement for healthcare professionals from the Stroke Council of the American Heart Association. Circulation. 2001;103(1):163–182.

10. Miller EL, Murray L, Richards L, et al. Comprehensive overview of nursing and interdisciplinary rehabilitation care of the stroke patient: a scientific statement from the American Heart Association. Stroke. 2010;41(10):2402–2448.

11. Foulkes MA, Wolf PA, Price TR, Mohr JP, Hier DB. The Stroke Data Bank: design, methods, and baseline characteristics. Stroke. 1988;19(5):547–554.

12. Rowe F, Brand D, Jackson CA, et al. Visual impairment following stroke: do stroke patients require vision assessment? Age Ageing. 2009;38(2):188–193.

13. Kapoor N, Ciuffreda KJ, Harris G, et al. A new portable clinical device for measuring egocentric localization. J Behav Optom. 2001;12(5):115–119.

14. Suchoff IB, Ciuffreda KJ. A primer for the optometric management of unilateral spatial inattention. Optometry-Journal of the American Optometric Association. 2004;75(5):305–318.

15. Chokron S, Imbert M. Variations of the egocentric reference among normal subjects and a patient with unilateral neglect. Neuropsychologia. 1995;33(6):703–711.

16. Keane S, Turner C, Sherrington C, Beard JR. Use of fresnel prism glasses to treat stroke patients with hemispatial neglect. Arch Phys Med Rehabil. 2006;87(12):1668–1672.

17. Heilman KM, Bowers D, Watson RT. Performance on hemispatial pointing task by patients with neglect syndrome. Neurology. 1983;33(5):661.

18. Padula W V, Nelson CA, Padula W V, Benabib R, Yilmaz T, Krevisky S. Modifying postural adaptation following a CVA through prismatic shift of visuo-spatial egocenter. Brain Inj. 2009;23(6):566–576.

19. Rossetti Y, Rode G, Pisella L, et al. Prism adaptation to a rightward optical deviation rehabilitates left hemispatial neglect. Nature. 1998;395(6698):166–169.

20. Houston KE. Measuring visual midline shift syndrome & disorders of spatial localization: a literature review & report of a new clinical protocol. Journal of Behavioral Optometry. 2010;21(4).

21. Facchin A, Beschin N, Daini R. Not prism prescription, but prism adaptation rehabilitates spatial neglect; a reply to Bansal, Han and Ciuffreda. Brain Inj. 2015;29(4):533–534.

22. Bansal S, Han E, Ciuffreda KJ. Use of yoked prisms in patients with acquired brain injury: A retrospective analysis. Brain Inj. 2014;28(11):1441–1446.

23. Kuhn C, Heywood CA, Kerkhoff G. Oblique spatial shifts of subjective visual straight ahead orientation in quadrantic visual field defects. Neuropsychologia. 2010;48(11):3205–3210.

24. Ferris FL, Kassoff A, Bresnick GH, Bailey I. New visual acuity charts for clinical research. Am J Ophthalmol. 1982;94(1):91–96.

25. neuroscience DPR neurology and, 2006 undefined. Postural disorders and spatial neglect in stroke patients: a strong association. https://content.iospress.com. Accessed March 14, 2023. https://content.iospress.com/articles/restorative-neurology-and-neuroscience/rnn00363

